# Theoretical Epidemic Laws Based on Data of COVID-19 Pandemic

**DOI:** 10.1101/2020.12.07.20238253

**Authors:** Junke Guo

## Abstract

The standard growth model of epidemic evolution such as the Richards generalized logistic function is remarkably successful because it agrees with almost all previous epidemic data. Yet, it fails to explain intervention measures for mitigations of the ongoing coronavirus 2019 disease (COVID-19) pandemic. It also fails to replicate an endemic phase that occurs in many countries epidemic curves (time series data of daily new cases). These discrepancies demonstrate that new epidemic laws are required to understand, predict and mitigate the COVID-19 pandemic. Here we show that almost all COVID-19 evolution can be modeled by three innovative epidemic laws. Specifically, based on the world COVID-19 data, we first divide an epidemic curve into three phases: an exponential growth phase, an exponential decay phase, and a constant endemic phase. We next integrate the growth and the decay phases into the first epidemic law with interventions as a model parameter. This law is completely opposite to the Richards generalized logistic function in terms of intervention measures. We then combine the first epidemic law with the endemic phase to form the second epidemic law, which makes the curve of cumulative cases increase linearly as time tends to infinity. The third epidemic law states if an epidemic is composed of multiple epidemic waves, the superposition principle applies. These laws were confirmed by the COVID-19 data from 18 countries including undeveloped, developing and developed countries. Finally, we pave the way for future research to incorporate the proposed theory into the classic SIR model. We anticipate that the results from this research can provide a scientific base for governments to mitigate the COVID-19 and other epidemic disasters.

## Introduction

### Problem and Significance

The Coronavirus 2019 disease (COVID-19) pandemic is an ongoing disease spreading all over the world. It has negatively impacted the world public health and economy. It caused more than 67M confirmed cases and more than 1.5M deaths since December 2019. It caused a world economic slowdown and made hundreds of millions of people unemployed. These numbers are continually climbing until herd immunity occurs naturally or a high percentage of the world population is vaccinated.

To suppress the spread of the COVID-19 pandemic before herd immunity, various mitigation measures have been implemented all over the world. Now, an important problem facing the world is: How do we assess the effects of various mitigation measures implemented to the COVID-19 pandemic and what did

we learn from it? Furthermore, How many people will be infected and how many people will die in this pandemic? When will this pandemic end? How does this pandemic affect the world economy? To answer these questions requires *epidemic laws* that accurately describe how an epidemic evolves under intervention measures. In this research, we discover epidemic laws based on the COVID-19 data facts.

### Data Facts

In epidemiology, a case is an individual infected by a disease; an epidemic curve is a curve of time series data of daily new cases; and a cumulative curve is a curve of total cases against time. Fig. 1(a) shows the cumulative curve of the first COVID-19 wave in Italy; and Fig. 1(b) is the corresponding epidemic curve. Mathematically, the change of the slope in Fig. 1(a) gives the epidemic curve in Fig. 1(b) that is observed with the following facts:

**Figure 1:**
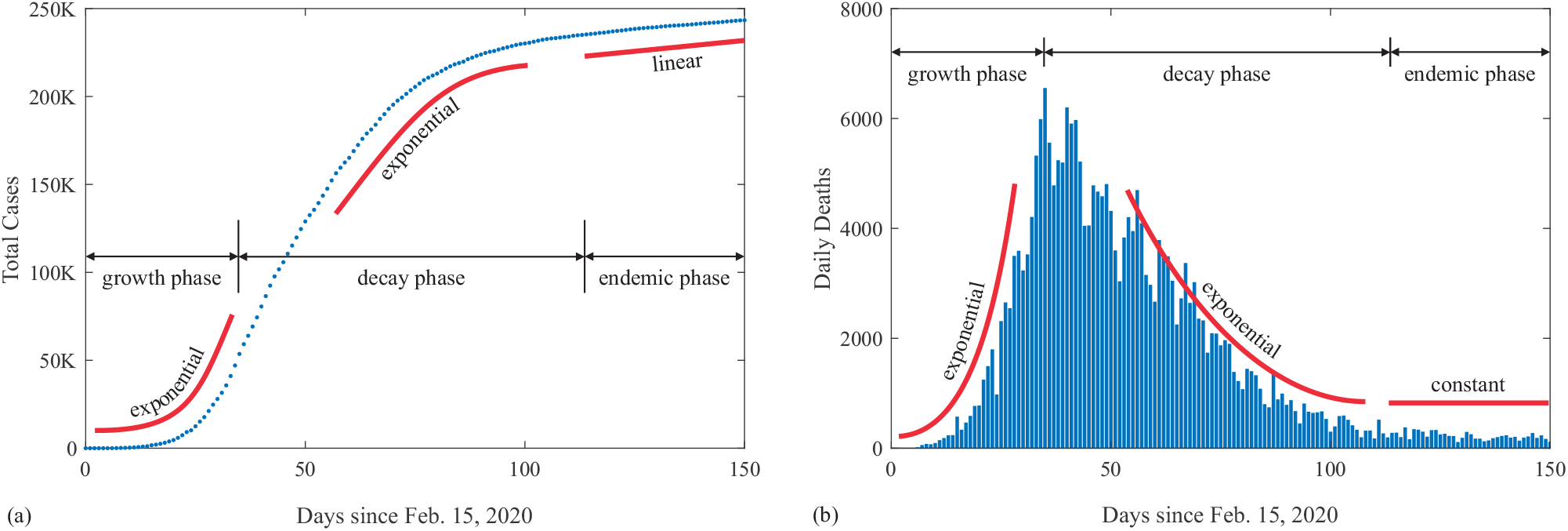
Data of the first COVID-19 wave in Italy (Worldometer 2020): (a) cumulative curve of total cases;(b) time series of daily new deaths.

- It has three phases: a growth phase, a decay phase, and an endemic phase.
- It increases exponentially in the growth phase.
- It decreases exponentially in the decay phase but the decay rate is affected by intervention measures.
- The growth and decay phases are connected with a hump but positively skewed.
- It tends to a constant asymptotically. If the constant is zero, the epidemic dies. Otherwise, the epidemic becomes an endemic that may trigger a second epidemic wave.

These facts make the cumulative curve in Fig. 1(a) bend upward exponentially in the growth phase and downward exponentially in the decay phase, and increase linearly in the endemic phase. These facts set the premises to justify an existing epidemic theory and to induce a new epidemic theory.

### Status and Gaps

Let us first justify that the current epidemic models or theories cannot replicate all of the facts in Figs. 1(a) and 1(b) accurately. For example, the most popular model to describe the cumulative cases against time in Fig. 1(a) is the Richards (1959) generalized logistic function:

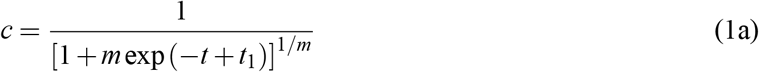

where *c* = dimensionless number of cumulative cases scaled by the carrying capacity; *t* = a dimensionless time; 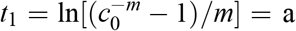 a dimensionless reference time with *c*_0_ as the initial condition; and *m* = an asymmetric parameter that is the only parameter to express the effects of intervention measures for mitigating the COVID-19 disaster. Thus, in this research, we call *m* the intervention parameter. At *m* → 0, Eq. (1a) reduces to the Gompertz (1825) function; and at *m* = 1, it reduces to the classic logistic function.

In terms of the initial condition, Eq. (1a) is rewritten as:

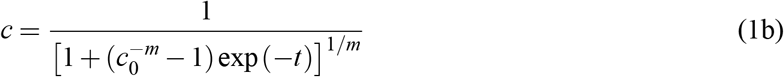

which is plotted in Fig. 2(a) with *m* ≥ 0 as a parameter and assuming *c*_0_ = 0.001. Let *t* → −∞ and consider *c*_0_ ≪ 1, Eq. (1b) has the left asymptote:

**Figure 2:**
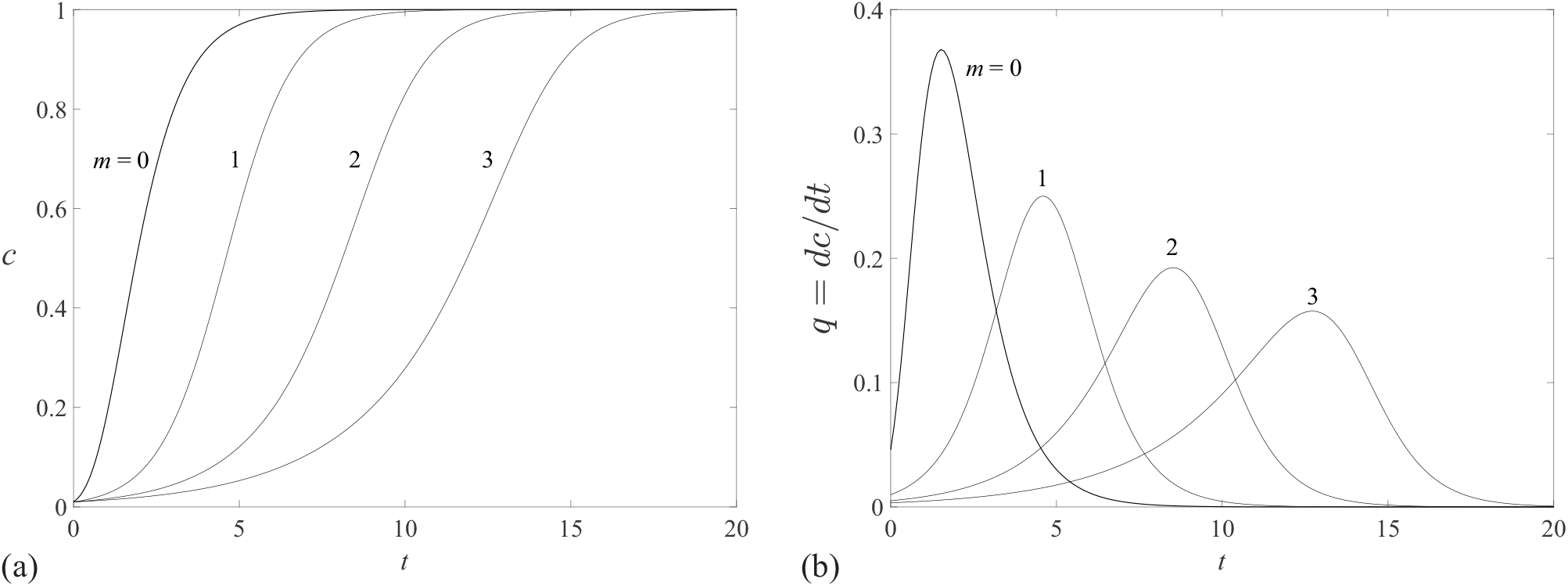
Generalized logistic function: (a) cumulative cuve from Eq. (1a); (b) daily new cases from Eq. (3)

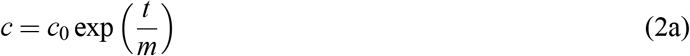

Similarly, let *t* → ∞ and consider *c*_0_ ≪ 1, Eq. (1b) has the right asymptote:

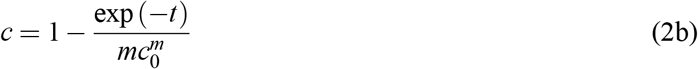

The dimensionless epidemic curve *q (t)* from Eq. (1b) is

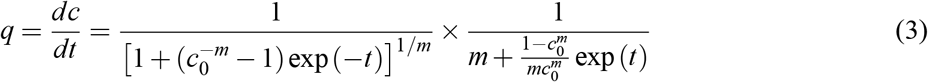

which is plotted in Fig. 2(b). Eq. (3) has the left asymptote:

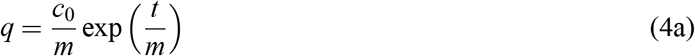

and the right asymptote:

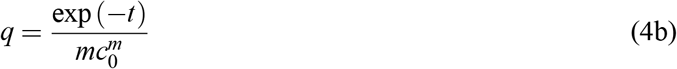

Eqs. (2a) and (4a) show that the intervention parameter *m* has a significant effect on the initial growth rate. Eqs. (2b) and (4b) show that in the decay phase, though the intervention parameter *m* has an effect on the magnitude, it has no effects on its decay rate. These conclusions contradict our common understanding of the topic that intervention mitigation measures do not affect the initial exponential growth but do significantly flatten the curve in the decay phase. Besides, as *t* → ∞, Eq. (2b) tends to 1 instead of a linear growth; and Eq. (4b) tends to zero instead of a positive constant. These results disagree with the data facts in the endemic phase in Figs. 1(a) and 1(b).

Therefore, in terms of the effects of intervention measures and the endemic phase, the Richards (1959) generalized logistic function fails to describe the COVID-19 evolution in all of the three phases. Besides, it provides little knowledge on an epidemic with multiple epidemic waves.

### Objective and Research Questions

The objective of this research is then to fill the above gaps with three innovative epidemic laws, which form a new epidemic theory. Specifically, we reach the objective by asking the following research questions:

1. Can we establish a theoretical law that correctly reflects the effects of intervention measures?
2. Can we find a simple epidemic law that accurately describes the evolution of COVID-19 that has an endemic phase?
3. How do we model an epidemic evolution with multiple epidemic waves?

### Hypotheses

We answer the three research questions by making the following hypotheses: A pandemic consists of many epidemics. An epidemic may have multiple epidemic waves. Referring to Fig. 1(b) and the data facts, an epidemic wave in terms of epidemic curves has the following features:

1. It increases exponentially in the growth phase.
2. It decreases exponentially in the decay phase but the decay rate is affected by intervention measures.
3. The growth and decay phases smoothly connect through a hump.
4. If an epidemic wave decays out, the epidemic curve approaches to zero as time tends to infinity.
5. If an epidemic wave decays to a positive constant as time tends to infinity, the epidemic becomes an endemic that may trigger a second epidemic wave.
6. If an epidemic has two or more epidemic waves, the superposition principle applies.

### Research Approach

We integrate the six hypotheses into a theory using both inductive and deductive reasoning:

*Step 1*: We integrate the first four hypotheses into a general differential equation. This process merges the two exponential laws in the growth and decay phases [Fig. 1(b)] into a single differential equation that results in the first epidemic law on cumulative cases without an endemic phase [Fig. 1(b)]. This law includes an explicit parameter expressing the effects of intervention mitigation measures, which answers our first research question. As a corollary of the first law, we deduce an epidemic curve equation on the time series of daily new cases.

*Step 2*: We generate the second epidemic law on cumulative cases by combining the first law with the endemic phase (Hypothesis 5) in Fig. 1(a). As a corollary of the second law, we deduce an epidemic curve equation that includes all three phases in Fig. 1(b). This step answers the second research question.

*Step 3:* We apply the superposition principle to an epidemic with multiple epidemic waves (Hypothesis 6) to form the third epidemic law. As a corollary of the third law, we deduce an expression to describe epidemic curves with multiple epidemic waves. This step results in the solution to the third research question.

*Step 4:* We test the three epidemic laws and their corollaries with the COVID-19 data that are from Worldometer (2020) at https://www.worldometers.info/coronavirus/.

### Epidemic Theory

Consider an isolated epidemic system with constant population *P*. At time *T*, the susceptible population is *S(T)*, and the infected population (or the number of cumulative cases) is *C(T)*. If the home population due to lockdowns is a constant *P*_*h*_. and the vaccinated population is a constant *P*_**v**_, then

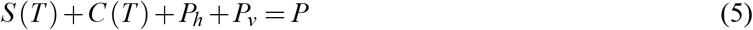

At *T*→ ∞, all susceptibles are infected, *S* (∞) = 0, the number of terminal cases from Eq. (5) is then

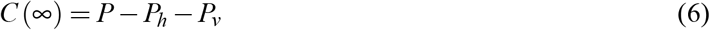

which states that to minimize the terminal cases *C*(∞), the best mitigation measure is to increase *P*_*v*_ by vaccinating a large part of the population. If without vaccines (*P*_*v*_ = 0), the second best measure is to let people stay home through lockdowns and thus to increase the home population *P*_*h*_. Here, we assume people are not susceptible if they stay home and do not contact with infected individuals.

Eqs. (5) and (6) result in the susceptible population at time *T* as:

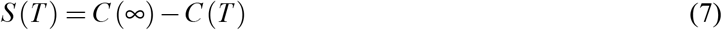

where the data of *C(T)* are reported everyday by health agencies (Worldometer 2020) as COVID-19 evolves. In what follows, we find three mathematical laws to describe *C(T)*, based on the COVID-19 data. For brevity, we denote *C*(∞)= *C*_1_.

### The First Epidemic Law without an Endemic Phase

Let *µ* = initial infection rate (d^−1^) that is the number of individuals infected by an existing case a day. This rate depends on virus features, climate conditions, and human behaviors, namely,

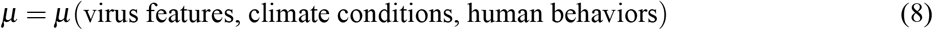

According to Hypothesis (1) and Fig. 1, the daily new cases *dC/dT* increases exponentially at *T*→ 0. Thus, the two numbers *C* and *dC/dT* in Figs. 1(a) and 1(b) satisfy the following differential equation:

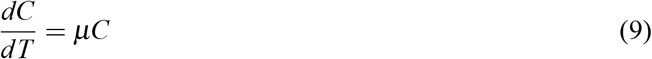

According to Hypothesis (2) and Fig. 1, in the decay phase, the daily new cases *dC/dT* decreases exponentially but the decay rate is affected by human interventions, which leads to

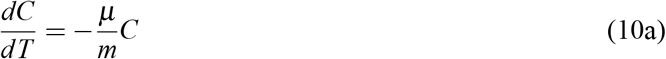

where similar to it in Eq. (1a), *m* = *intervention parameter* expressing change of human behaviors in Eq. (8). Hypothesis (4) requires *dC/dT* = 0 at *T*→ ∞. Considering *C*(∞) = *C*_1_, we revise Eq. (10a) as:

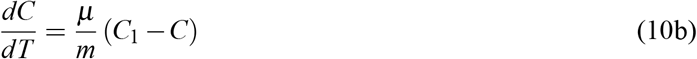

where *µ/m* = effective infection rate with intervention measures = decay rate, and *C*_1_−*C* = susceptible at time *T* according to Eq. (7). Eq. (10b) states that the most efficient way to suppress the daily new cases *dC/dT* is to minimize the susceptible *C*_1_−*C* by reducing *C*_1_ through Eq. (6). This is what China did in controlling the COVID-19 virus despite vaccines. The second way is to make the intervention parameter *m* > 1 by changing human behaviors such as keeping social distancing, mask wearing, and hand hygiene, which reduces the effective infection rate *µ/m* and thus reduces the daily new cases *dC/dT*. The second way is widely practiced in the rest of the world.

Taking Eqs. (9) and (10b) as premises, by analogy to an eddy viscosity model for turbulent open channel flow (Guo 1988), we induce the following generalized differential equation:

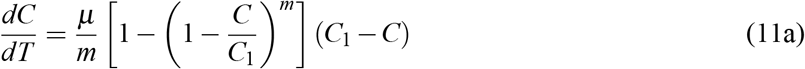

where 1 −(1 *C*−/*C*_1_)^*m*^ = probability that susceptible meet infected individuals, which is intuitive if taking *m* = 1 (natural spread) and assuming *P*_*h*_ = *P*_*v*_ = 0 in Eqs. (5) and (6). Eq. (11a) is then interpreted from the right to the left side as: At time *T*, the system has susceptible *C*_1_−*C* who meet infected individuals with a probability 1−(1−*C/C*_1_)^*m*^. If the effective infection rate is *µ/m*, then the product of the three factors is the daily new cases *dC/dT*.

Defining *c* = *C/C*_1_ as the dimensionless number of *C* and *t* = *µ T* as the dimensionless number of *T*, Eq. (11a) reduces to

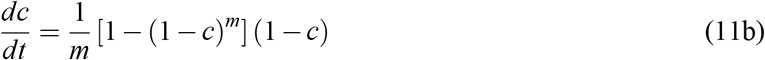

which has three special cases:

(1) At *m* → 0, according to L’Hopital’s rule, lim_*m*→0_[1 − (1 −c)^*m*^]/*m* = −ln(1−*c*), which makes Eq. (11b) grows fastest, namely,

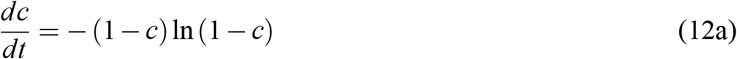

which results in a double exponential law:

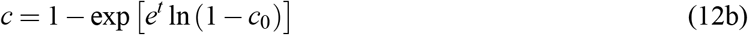

where *c*_0_ = the dimensionless initial condition at *t* = 0. This case represents the fastest growth of a disease spread and is plotted in Fig. 3, assuming *c*_0_ = 0.05.

**Figure 3:**
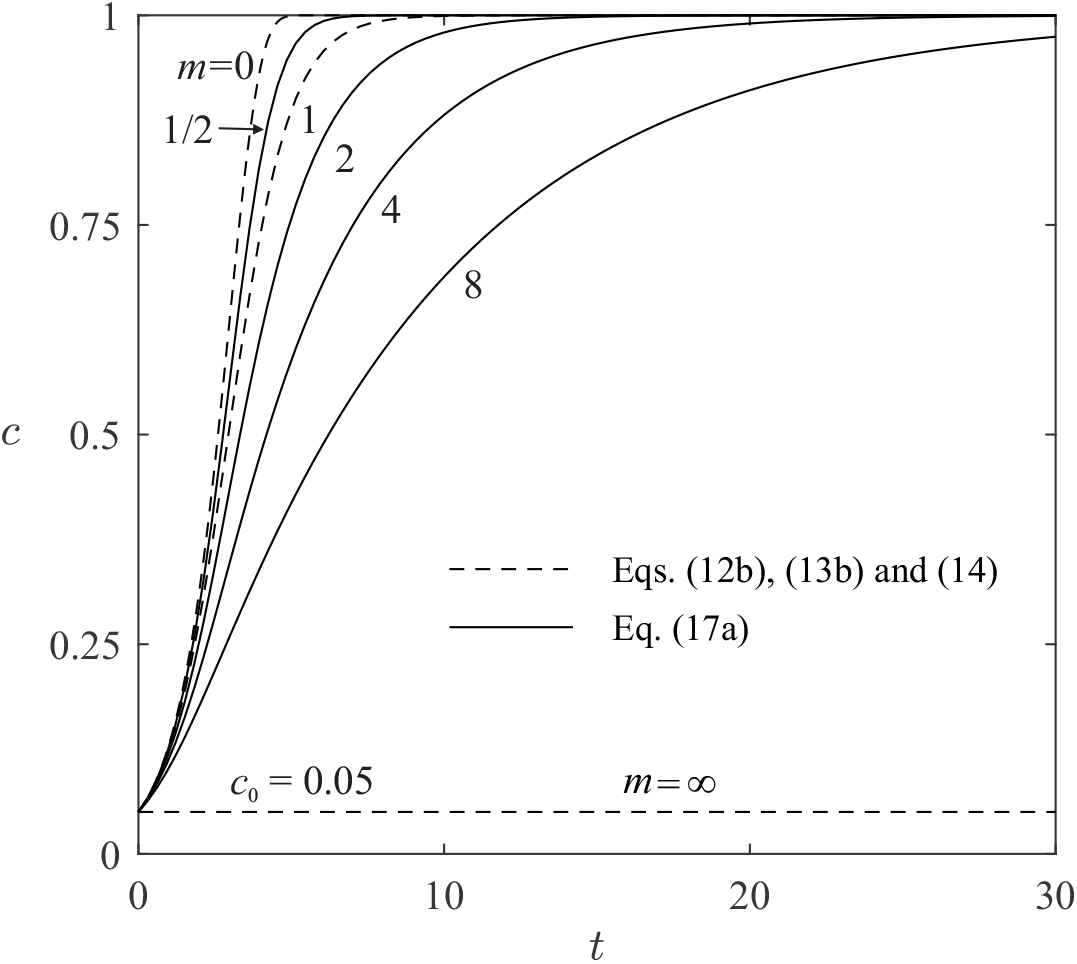
Effects of intervention parameter *m* in the number of cumulative cases from Eq. (17a)

(2) At *m* = 1, Eq. (11b) reduces to

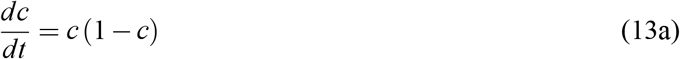

which results in the standard logistic function:

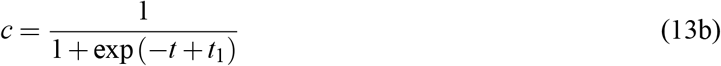

where *t*_1_ = dimensionless inflection time at *c* = 1/2. Eq. (13b) is plotted in Fig. 3 and represents a natural spread of diseases under constant environmental and societal conditions.

(3) At *m* → ∞, Eq. (11b) reduces to: *dc/dt* = 0, which gives

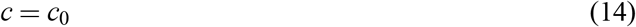

plotted in Fig. 3. This extreme case states we could have suppressed COVID-19 to its initial condition *c*_0_ and squashed it in the bud if we had taken extreme mitigation measures (*m* → ∞), for example, suspending all international airlines and ships worldwide in February 2020.

On proceeding to the general solution, let

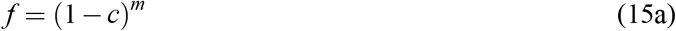

or

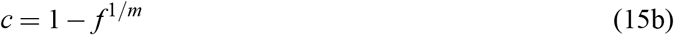

then Eq. (11b) reduces to

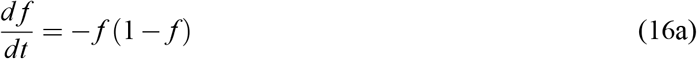

resulting in

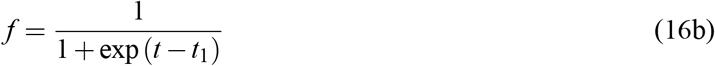

where *t*_1_ = dimensionless inflection time at *f* = 1/2. Because Eqs. (13b) and (16b) are a pair of complementary functions and *t*_1_ is their cross point, *t*_1_ in Eq. (16b) is then the dimensionless natural inflection time. Eq. (16b) is the Fermi function widely used in quantum mechanics (Fermi et al. 1955).

Inserting Eq. (16b) into Eq. (15b) gives

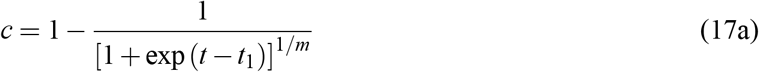

where the natural inflection time *t*_1_ is determined by the initial condition, *c*(0)= *c*_0_, namely,

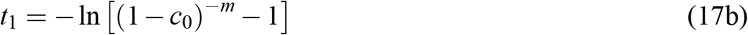

By analogy to the Richards (1959) generalized logistic function in Eq. (1a), we call the fraction in Eq. (17a) a generalized Fermi function. Hence, Eq. (17a) is the complement of the generalized Fermi function.

Let *t* → − ∞ and consider *c*_0_ ≪ 1, Eq. (17a) reduces to:

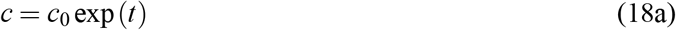

which shows that intervention measures have no effects on the initial exponential growth. Let *t* → ∞ and consider *c*_0_ ≪ 1, Eq. (17a) reduces to

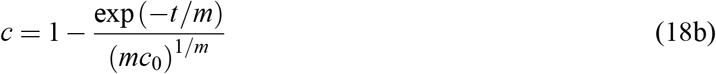

which states that intervention measures change both the magnitude and the decay rate in the decay phase. These extremes agree with our common understanding of the topic but are completely opposite to Eqs. (2a) and (2b) from the Richards (1959) generalized logistic function.

The effects of the intervention parameter *m* are further explained in Fig. 3 by plotting Eq. (17a). It shows that: as 0≤ *m*< 1, the cumulative curve *c(t)* is raised, compared with the natural spread (*m* = 1); as *m* increases from 1, the cumulative curve *c(t)* is flattened; as *m*→ ∞, the disease is completely squashed to its initial condition *c*_0_. Mathematically, Eq. (17a) contains the three special cases so we call it the first epidemic law, which answers our first research question.

#### Corollary 1

The dimensionless epidemic curve *q(t)* in terms of the daily new cases is derived by inserting Eq. (15b) into Eq. (11b), resulting in

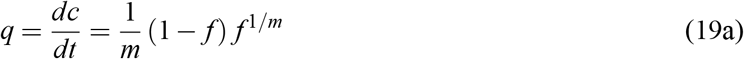

Applying *f* from Eq. (16b) in Eq. (19a) gives

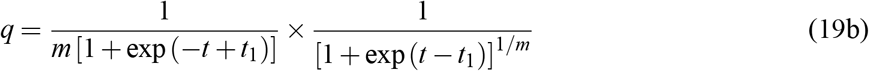

Assuming *c*_0_ = 0.001 in Eq. (17b) for *t*_1_, Eq. (19b) is then plotted in Fig. 4(a) in linear scale and Fig. 4(b) in logarithmic scale. Letting *dq/dt* = 0 gives the maximum of daily cases at

**Figure 4:**
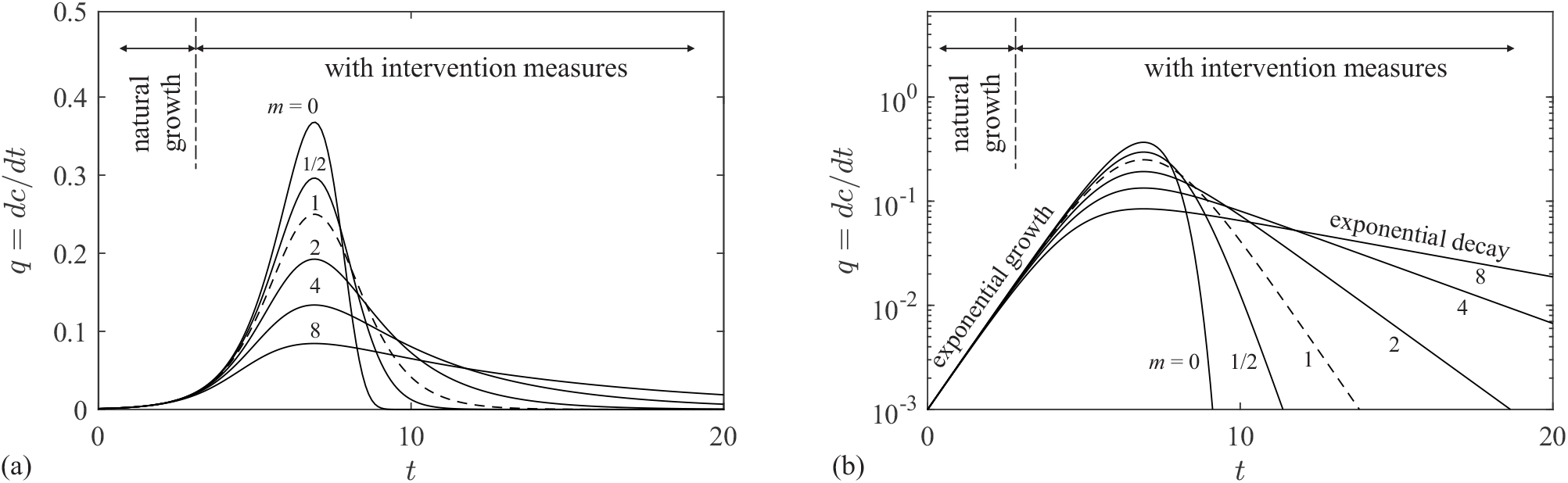
Dimensionless epidemic curves in *q(t)* from Eq. (19b), assuming *c*_0_ = 0.001: (a) linear scale; and (b) logarithmic scale.

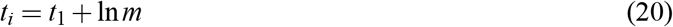

which states that the real inflection time *t*_*i*_ in Fig. 3 is postponed by ln *m* if mitigation measures are applied (*m* > 1). If *m*< 1, the real inflection time *t*_*i*_ appears earlier than the natural inflection time *t*_1_.

If *m* = 0, Eq. (12b) yields

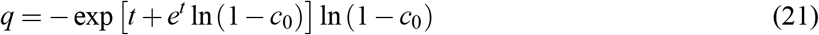

which gives both the fastest growth and the fastest decay of daily new cases, as shown in Fig. 4.

If *m* = 1, Eq. (19b) reduces to

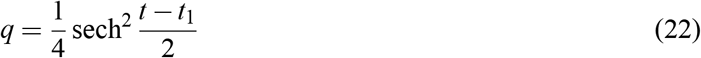

which is the symmetric solitary wave equation in shallow water hydrodynamics (Korteweg and de Vries 1895), quantum mechanics (Fermi et al. 1955) and mathematics (Wazwaz 2009). Thus, in this research, we call a single epidemic wave a solitary epidemic wave or simply an epidemic soliton.

If 0≤ *m* < 1, the epidemic curves from Eq. (19b) grow and decay faster than the natural disease spread (m = 1), as shown in Fig. 4(a). This states we can quickly reach herd immunity and end an epidemic by mixing susceptible and infected individuals by mass gatherings and events. Nevertheless, this may quickly overwhelm public health systems and thus risk population lives. To save lives from COVID-19 before vaccines are invented, we should flatten the curves in Fig. 4(a) by increasing *m* from 1.

Particularly, Fig. 4(a) shows that unlike Fig. 2(b) from the generalized logistic function, the intervention parameter *m* does not affect the initial growth, as expected from Eq. (18a). This is also explained in Fig. 4(b) with the logarithmic scale. Furthermore, Fig. 4(b) shows that Eq. (19b) successfully replicates the two exponential laws in the growth and decay phases. Unfortunately, similar to the Richards (1959) generalized logistic function of Eq. (1a), Eq. (19b) gives *q*(*∞*) = 0 and thus it cannot replicate the endemic phase in Fig. 1(a), which is addressed below.

### The Second Epidemic Law with an Endemic Phase

The endemic phase is an equilibrium state of an epidemic system under constant environmental and societal conditions. If these conditions are disturbed by changes of climate, human behaviors and interventions, the endemic triggers a second epidemic wave. Therefore, the endemic phase should be included if an epidemic does not die asymptotically.

According to Hypothesis (5) and Fig. 1(b), an endemic phase exists if *dc/dt* = *a* at *t* → ∞, where *a* = a small positive constant. In terms of cumulative cases (integration), this implies at *t* → ∞, we have

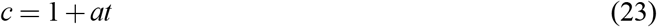

where the integration constant 1 is obtained from the first epidemic law where *c*(∞) = 1 at *a* = 0.

We can generalize Eq. (23) and the first epidemic law of Eq. (15b) into a single equation by *c* = Eq. (15b) × Eq. (23), resulting in

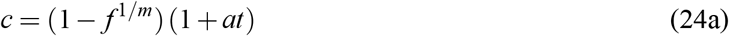

where (1 + *at*) = a dimensionless endemic factor. Applying Eq. (16b) in Eq. (24a) gives

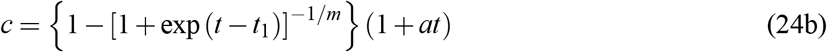

which is plotted in Fig. 5(a), assuming *c*_0_ = 0.001 and *m* = 4. Eq. (24a) or (24b) is **the second epidemic law** describing the cumulative cases with an endemic phase.

**Figure 5:**
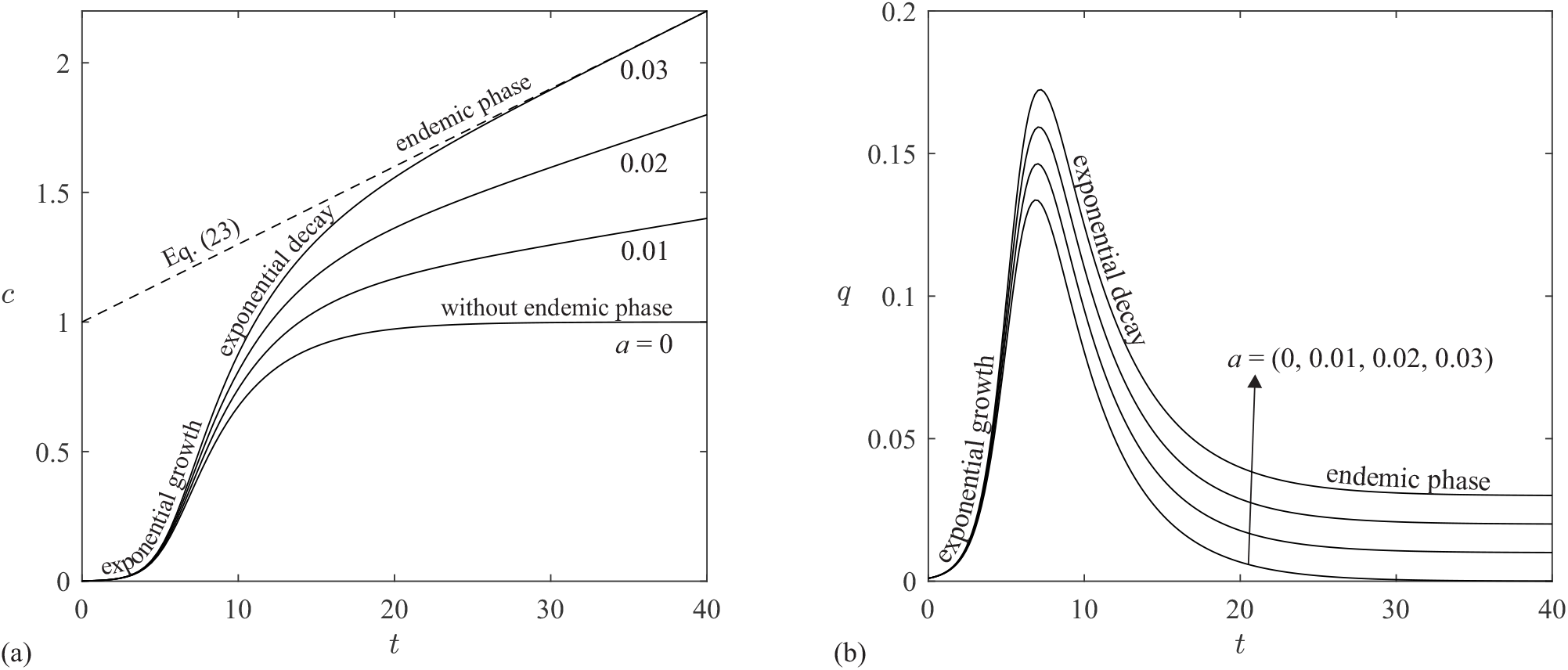
Graphical interpretation of the second epidemic law: (a) dimensionless cumulative cases from Eq. (24b); (b) epidemic curve from Eq. (25b), assuming *c*_0_ = 0.001 and *m* = 4

Similar to the intervention parameter *m*, the endemic constant *a* has no effects on the initial growth, as shown in Fig. 5(a). Besides, if *a* = 0, Eq. (24b) reduces to Eq. (17a) so the first epidemic law is a special case of the second epidemic law.

#### Corollary 2

The dimensionless epidemic curve from Eq. (24a) is derived as follows:

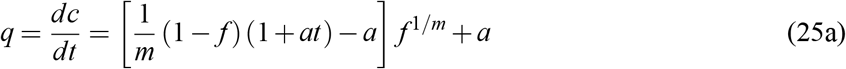

or

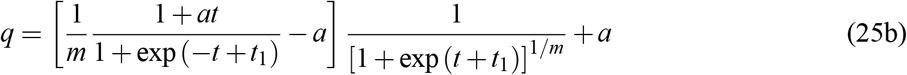

which is plotted in Fig. 5(b) on the effects of the endemic constant *a*, assuming *c*_0_ = 0.001 and *m* = 4. Clearly, if *a* = 0, the number of daily cases *q* tends to zero at *t* → ∞, implying that an epidemic eventually dies; if *a* is very small, the epidemic eventually becomes an endemic and it is controllable; yet, if *a* is very large, the epidemic is out of control and many epidemic waves are coming one after another.

Let *dq/dt* = 0 in Eq. (25a), we obtain that the maximum of daily cases in Fig. 5(b) occurs at

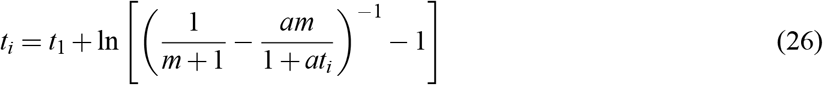

which is an implicit equation for *t*_*i*_. Eq. (26) states that the real inflection time *t*_*i*_ is further postponed if an endemic exists, as shown in Fig. 5(b).

Note that for *a* > 0, the value of *C*_1_ is simply a scale of *C* because an asymptotic value of *C* does not exists in Fig. 5(a). Fig. 5(b) shows that Eq. (25a) replicates all of the three phases in Fig. 1(b), which provides an advantage over the Richards (1959) generalized logistic function, including the Gompertz (1825) function and the classic logistic function. Therefore, the second epidemic law of Eq. (24a) and Corollary 2 of Eq. (25a) answer our second research question.

### The Third Epidemic Law for Multiple Epidemic Waves

An epidemic system may have multiple epidemic waves because of several reasons: (1) It may have multiple disease sources; (2) the endemic phase is disturbed by changes of mitigation measures or other factors; and (3) the case scale *C*_1_ or *C* (∞) may not be constant in Eq. (6) because of national holidays, celebrations, elections, protestations, and so on.

Once multiple epidemic waves occur, Hypothesis (6) states that the superposition principle applies. First, if herd immunity is achieved after many epidemic waves, then *q* (∞) = 0, which requires *a* = 0 in Eqs. (24a) and (25a). Therefore, each wave is an independent solitary wave described by the first epidemic law. The superposition principle in terms of dimensional form then gives

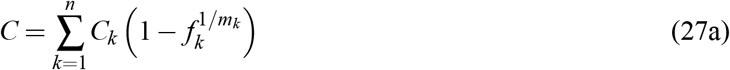

where *C*_*k*_ and *m*_*k*_ = scale of cases and intervention parameter for the *k*-th epidemic wave, respectively; and the corresponding Fermi function from Eq. (16b) is

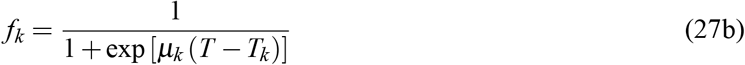

where *µ*_*k*_ and *T*_*k*_ = initial infection rate and natural inflection time for the *k*-th wave, respectively.

Nevertheless, if an endemic exists even after n waves (Fig. 6), then as *t* → ∞, all waves share a common dimensionless endemic factor (1 + *at*) or (1 + *AT*) with *A* as a small dimensional endemic constant (d^−1^). Therefore, the superposition from the second epidemic law gives

**Figure 6:**
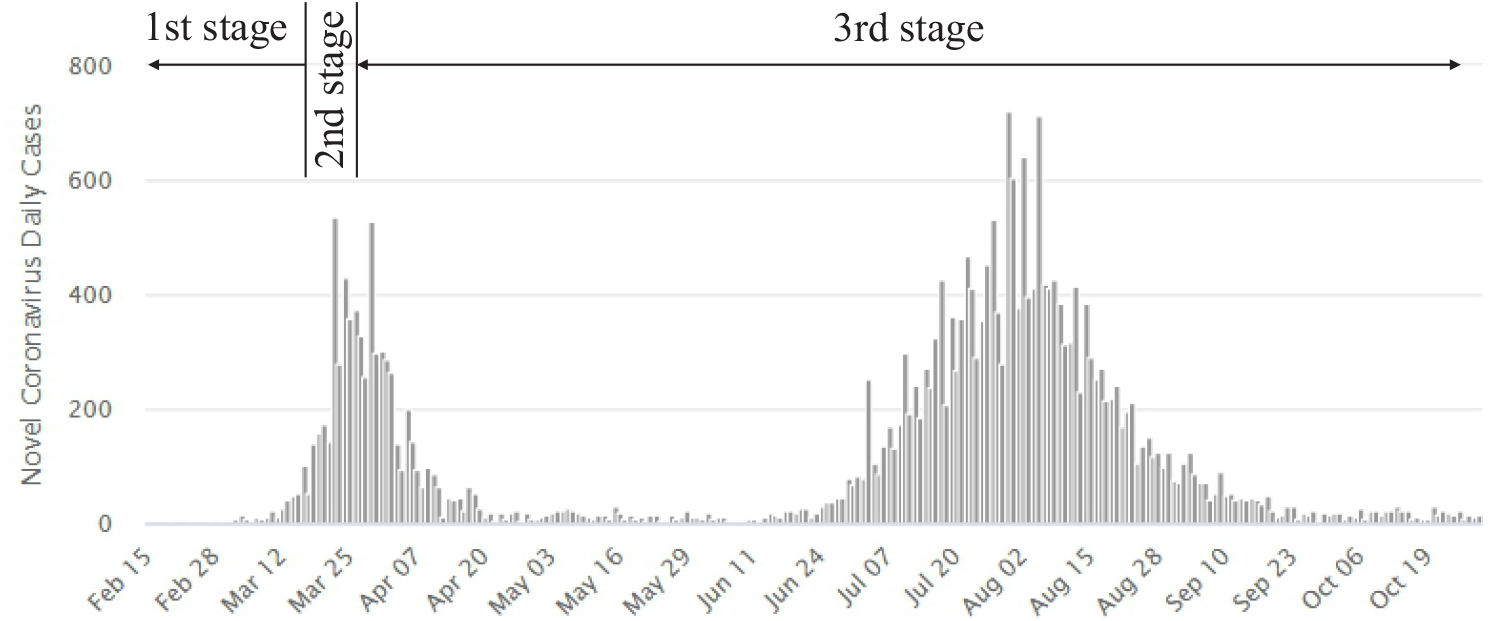
Epidemic curve of COVID-19 in Australia with multiple waves (Worldometer 2020)

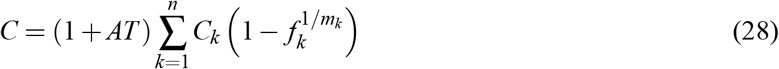

which is the **third epidemic law** describing the cumulative cases with multiple epidemic waves. Eq. (28) answers our third research question.

#### Corollary 3

Taking the derivative of Eq. (28) with respect to *T* gives the daily new cases

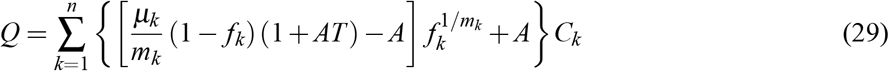

As *T* → ∞, Eq. (29) yields

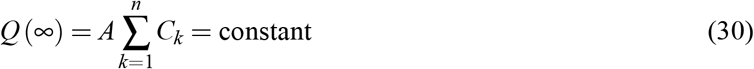

which has three possible results: (1) If *Q*(∞) is zero, then the epidemic dies or herd immunity is achieved, which corresponds to Eq. (27a); (2) if *Q*(∞) is small, the epidemic is controllable even without vaccines; and (3) if *Q* (∞) is large, then the epidemic transmission is out of control and the only way to suppress Eq. (30) is to vaccinate a large part of population and thus to reduce the susceptible *C*_*k*_ according to Eq. (6).

### Test with COVID-19 Data

#### Population and Sample Data

The three epidemic laws are derived for the cumulative cases from the population data of a closed system (state or country). Nevertheless, the real number of cumulative cases is never known if without testing of the entire population. All we know is the number of cumulative confirmed cases and the cumulative deaths. In this research, we assume (1) the sample dataset is similar to the population dataset,

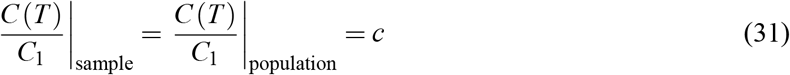

(2) the number of cumulative deaths (D) is also described by the three laws because of similarity of time series data between daily new cases and daily deaths (Worldometer 2020).

### Determination of Model Parameters

For a solitary epidemic wave, the second epidemic law of Eq. (24b) has five model parameters: the infection rate *µ* the scale *C*_1_ for cases; the intervention parameter *m*; the natural inflection time *T*_1_; and the endemic constant *A*. They are obtained by fitting Eq. (24b) to data in terms of *C* (*T*), using the Matlab (2020b) least-squares function lsqcurvefit.m. The initial values of these parameters are set as follows:

- The infection rate *µ*, defined by Eq. (9), is the daily new cases produced by an existing case during an incubation period. We assume once a case is identified, it is completely contained without transmission. According to WHO (2020), the incubation period of COVID-19 is between 2 and 14 days and the reproductive number *R*_0_ is between 1.4 and 2.5. This research then assumes *R*_0_ = 2 and the incubation period to be 10 days in such a way that we set *µ* ≃ 0.2 d ^1^.
- The scale *C*_1_ starts at *C*_1_ = max (*C*). For death curves, *C*_1_ is replaced by *D*_1_.
- The intervention parameter *m* starts at *m* = 4 if mitigation measures are applied.
- The natural inflection time *T*_1_ starts at *T*_1_ = *T*_*i*_ corresponding to the maximum of daily cases.
- The value of A starts at *A* = 0d^−1^ because *A* ≥ 0.

Similar initial values are set for each epidemic wave if an epidemic consists of multiple waves.

### Test of the Second Epidemic Law with Data of Solitary Epidemic Waves

The first epidemic law is a special case of the second law that is best tested with data from countries that are still in the first wave but with a fully developed endemic phase, as shown in Fig. 7 where (*C, Q*) = cumulative cases and daily cases in sub-figures (a) and (b), respectively, and (*D, Q*_*d*_) = cumulative deaths and daily deaths in sub-figures (c) and (d), respectively. Fig. 7 includes data from 11 countries, including South Africa, Chile, Bolivia, Qatar, Egypt, Eswatini, French Guyana, Gambia, Malawi, Mauritania, and Yemen. These data confirm the second epidemic law and its corollary. Fig. 8 further validates the second law with the first wave data from six developed countries, including the U.K., Germany, France, Spain, Italy, and the State of New York in the U.S.

**Figure 7:**
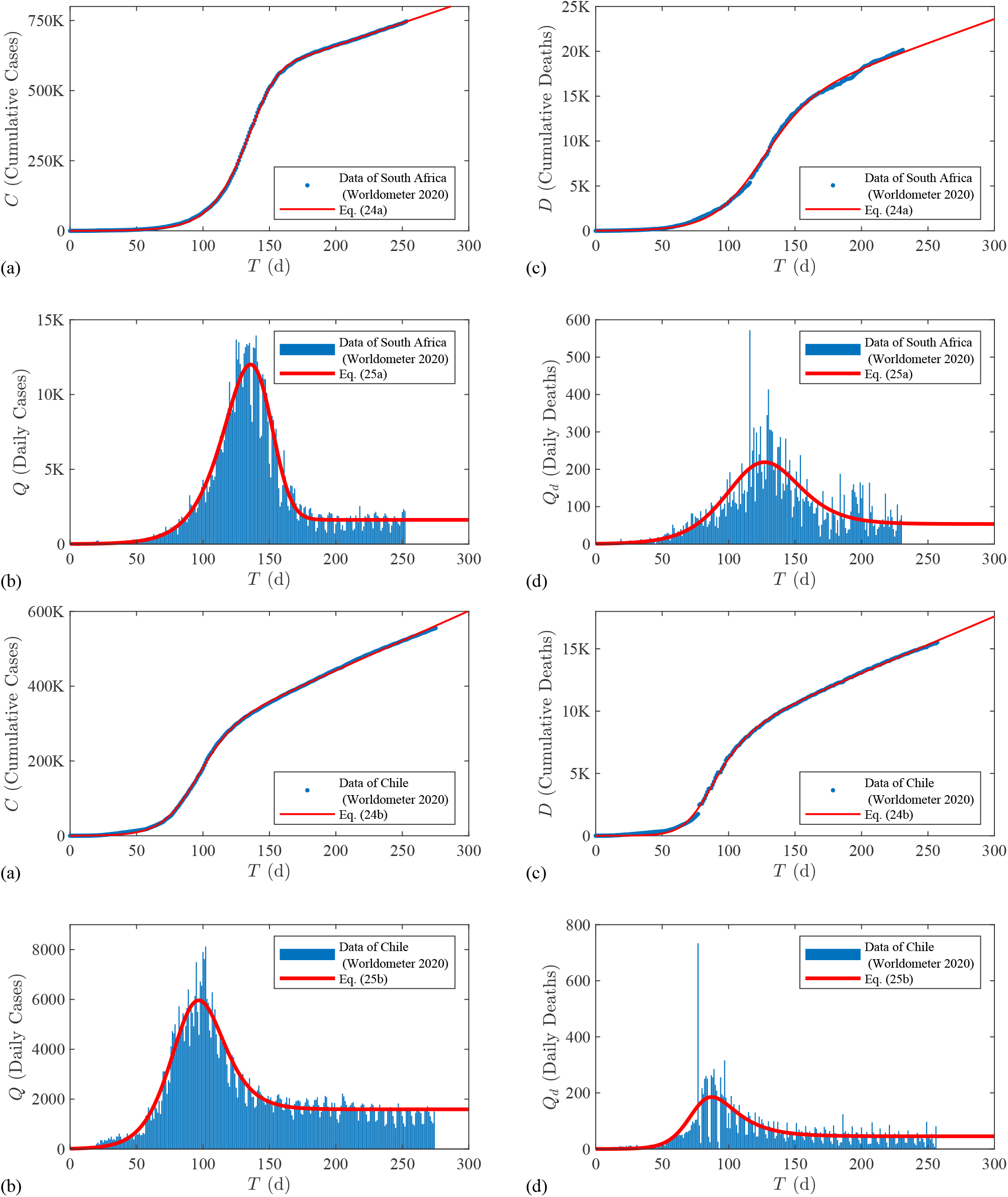
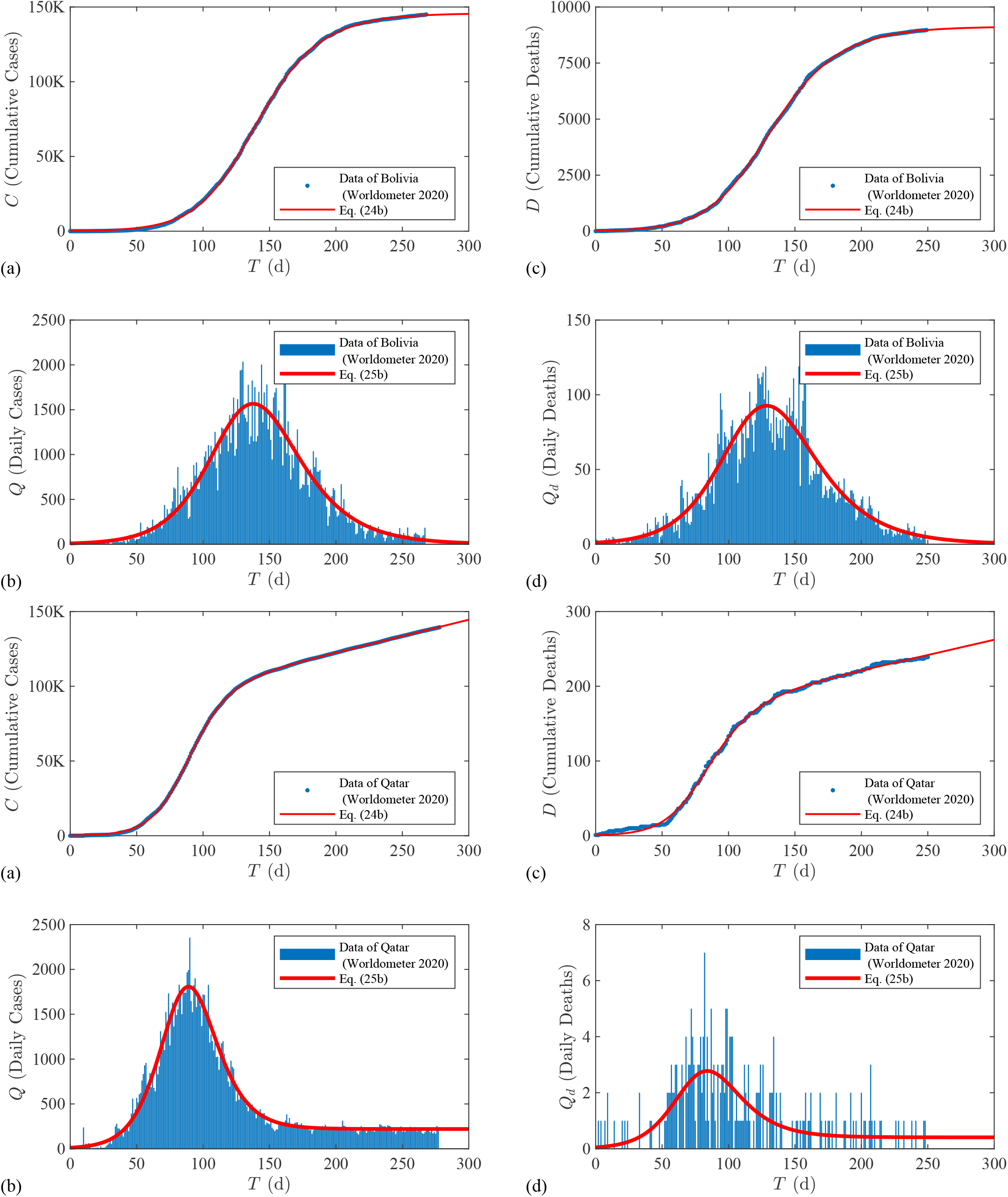
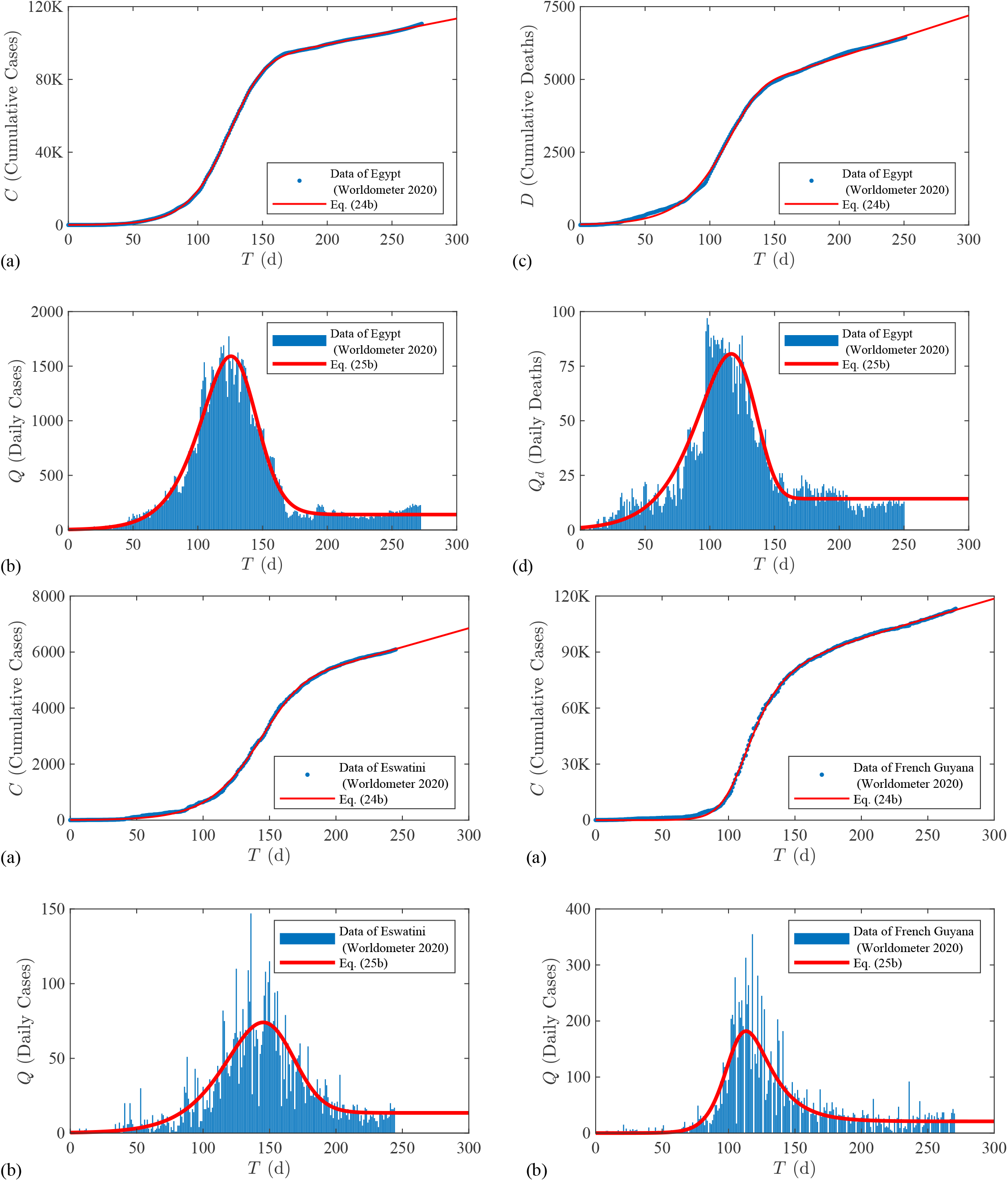
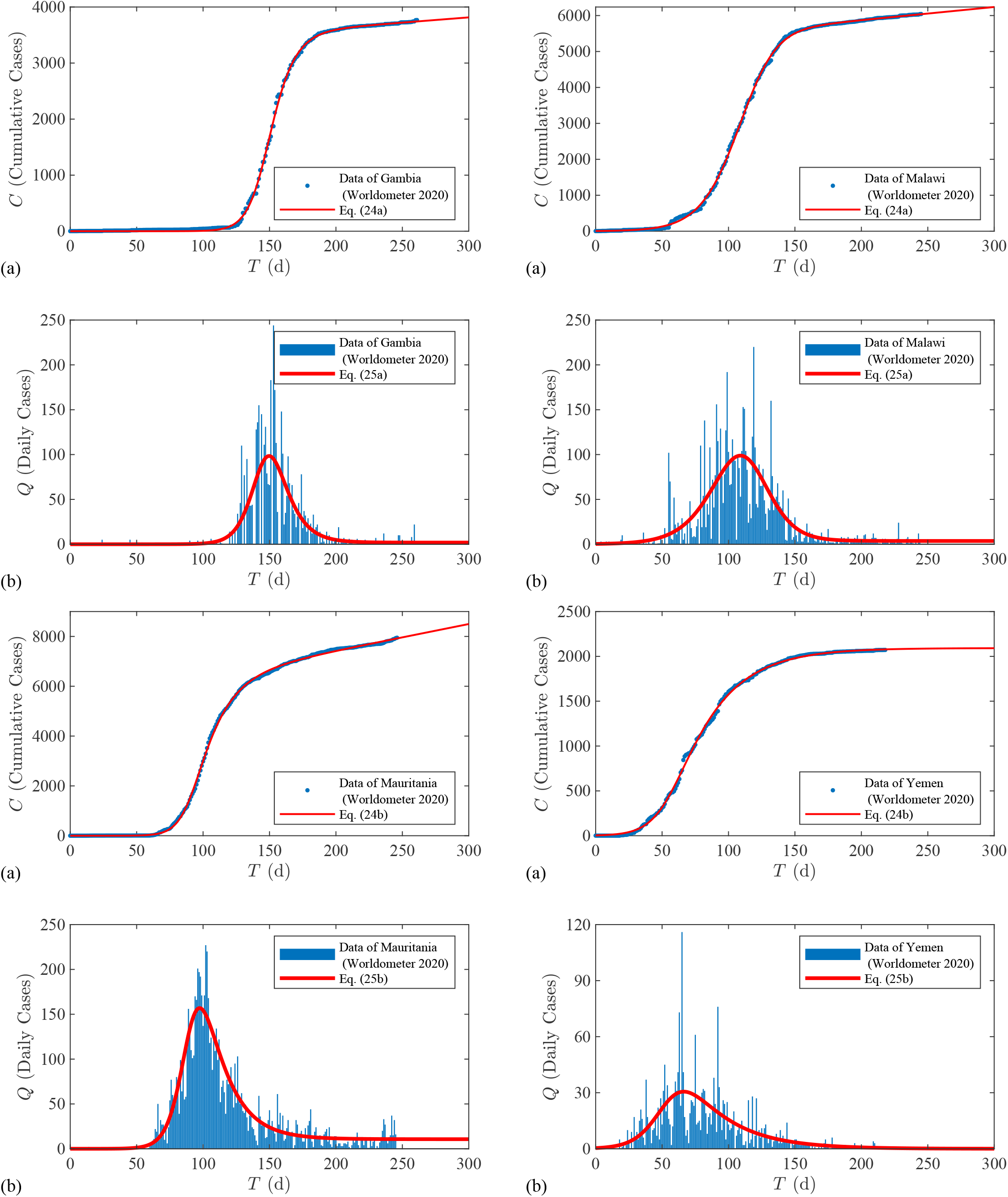
Test of the second epidemic law with data from 11 countries that are in the first waves with a developed endemic phase.

**Figure 8:**
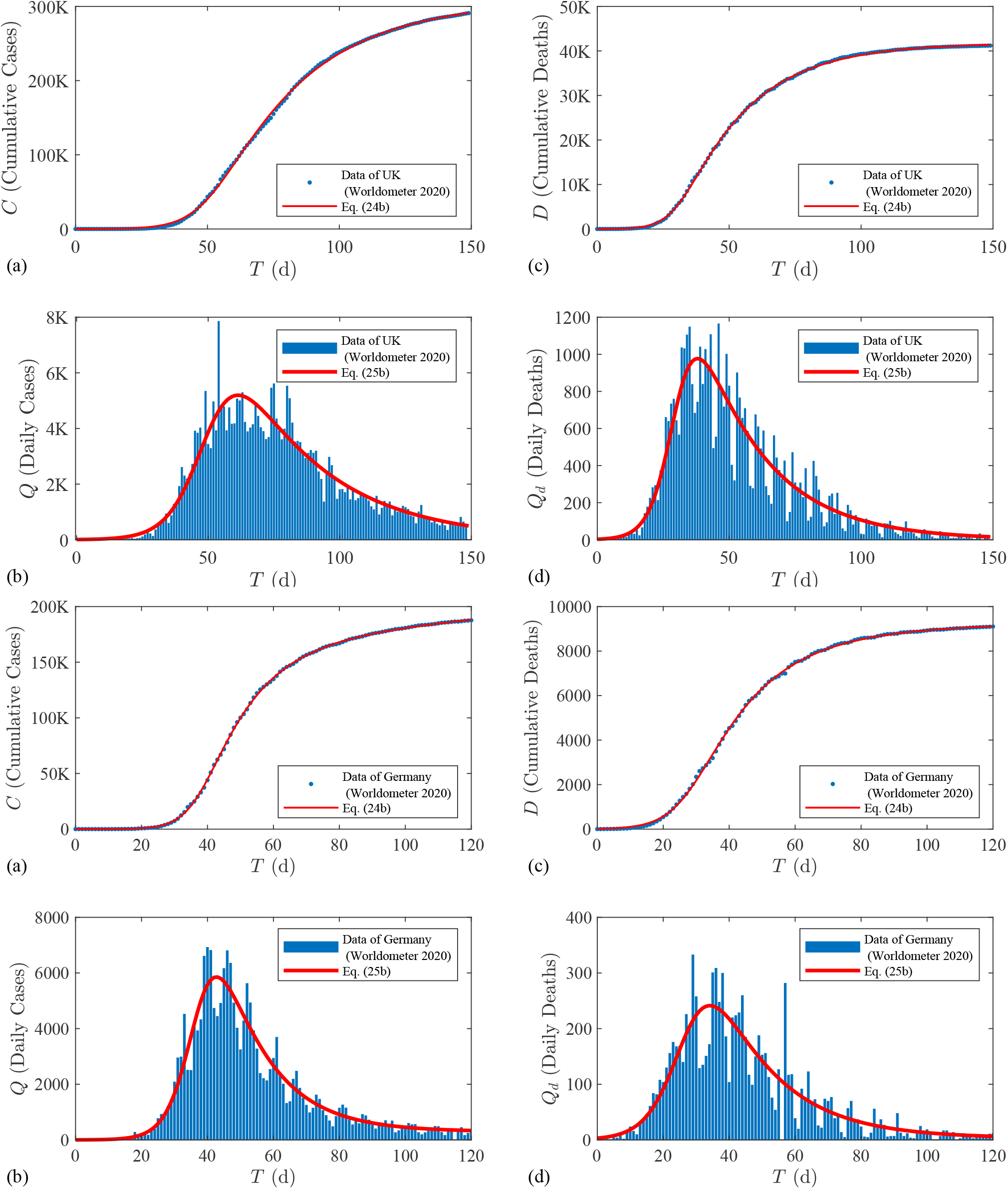
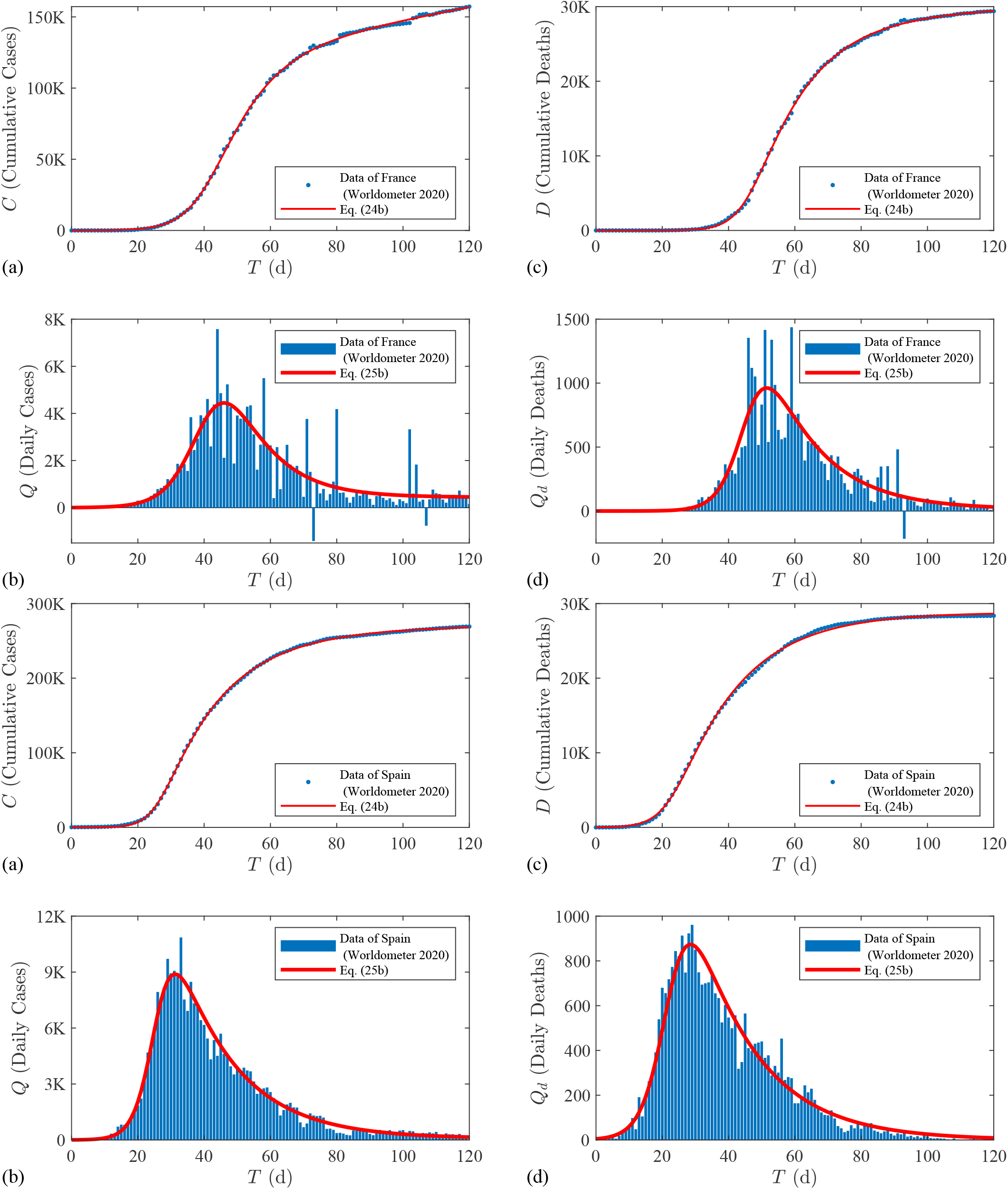
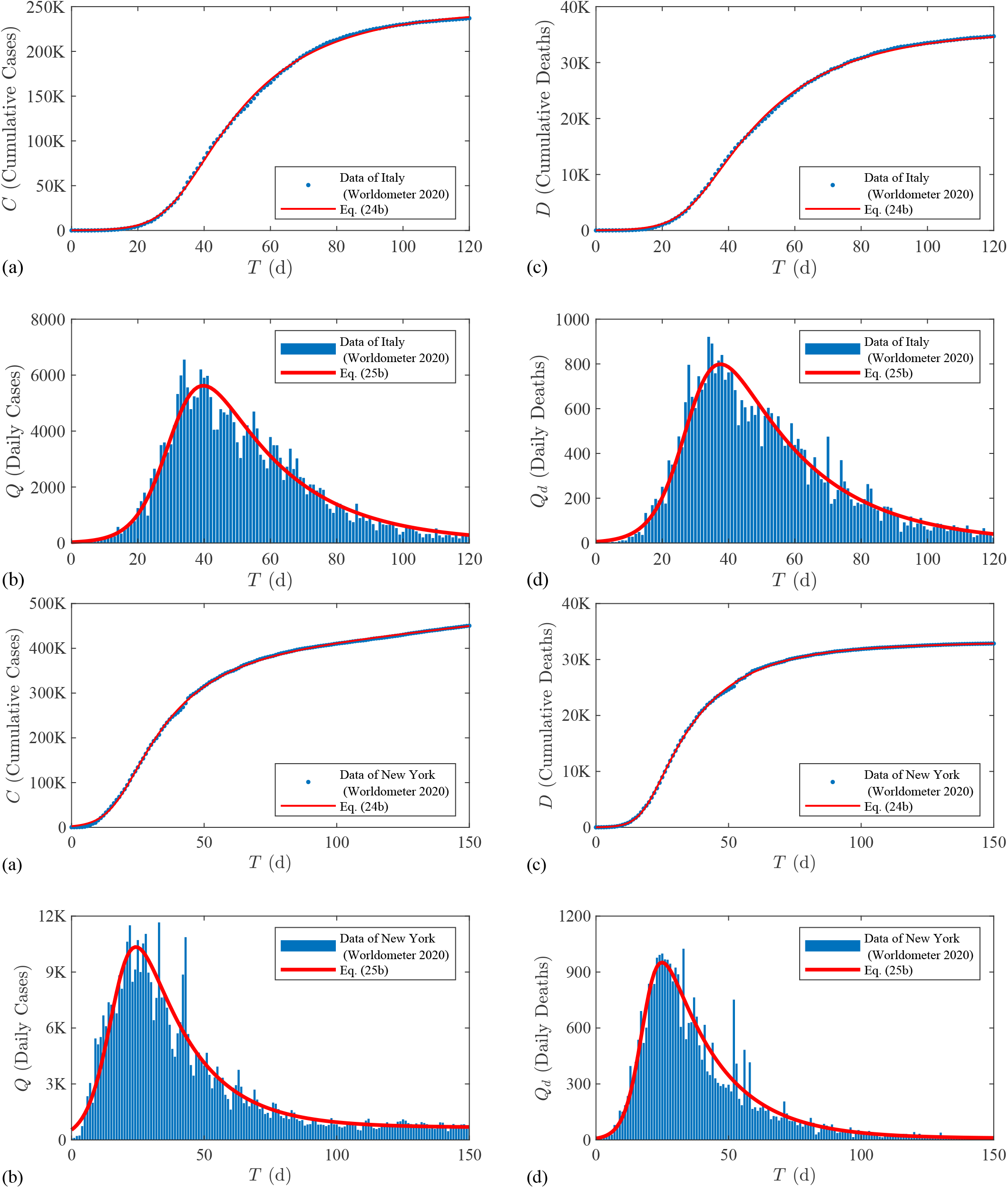
Test of the second epidemic law and its corollary with the first COVID-19 wave data from in the U.K., Germany, France, Spanish, Italy, and the State of New York in the U.S.

The model parameters from Figs. 7 and 8 are tabulated in Table 1. It shows that: (1) The mean value of the initial infection rate, *µ* = 0.08 d^−1^, from Fig. 7 is much less than *µ* = 0.21 d^−1^ from Fig. 8. The mean value of the natural inflection time, *T*_1_ = 118 d, from Fig. 7 is much larger than *T*_1_ = 34 d from Fig. 8. These values mean that COVID-19 spreads much faster in developed countries because of active economics. (2) The mean value of the intervention parameter *m* for Fig. 7 is close to 1, meaning that COVID-19 spreads in some undeveloped and developing countries almost naturally without mitigations. Yet, the mean value of *m* is 4 for Fig. 8, meaning that COVID-19 is significantly suppressed by mitigation measures in developed countries. (3) Although the endemic constants A in Table 1 for cases are small, they are the flames to trigger second waves. (4) The death parameters are comparable with the case parameters because the death number increases with increasing the case number.

**Table 1:** Epidemic parameters for data in Figs. 7 and 8

| Cases |  |  |  |  |  | Deaths |  |  |  |  |
| --- | --- | --- | --- | --- | --- | --- | --- | --- | --- | --- |
| Country | $\mu$<br>(d<sup>-1</sup>) | $T_1$<br>(d) | $C_1$ | $m$ | $A$<br>(d<sup>-1</sup>) | $\mu$<br>(d<sup>-1</sup>) | $T_1$<br>(d) | $D_1$ | $m$ | $A$<br>(d<sup>-1</sup>) |
| South Africa | 0.059 | 162 | 336779 | 0.19 | 0.0048 | 0.051 | 125 | 7505 | 0.85 | 0.00715 |
| Chile | 0.068 | 99 | 110675 | 0.69 | 0.015 | 0.098 | 81 | 3531 | 1.49 | 0.0135 |
| Bolivia | 0.048 | 131 | 145757 | 1.38 | 3.4e-11 | 0.046 | 120 | 9021 | 1.45 | 5.16e-05 |
| Qatar | 0.071 | 86 | 78234 | 1.08 | 0.00283 | 0.061 | 82 | 127 | 0.98 | 0.00373 |
| Egypt | 0.055 | 141 | 71097 | 0.39 | 0.00198 | 0.043 | 194 | 2905 | 0.03 | 0.00492 |
| Eswatini | 0.043 | 172 | 2799 | 0.27 | 0.00482 |  |  |  |  |  |
| French Guiana | 0.112 | 106 | 5697 | 1.98 | 0.00358 |  |  |  |  |  |
| Gambia | 0.126 | 146 | 3314 | 1.48 | 0.000482 |  |  |  |  |  |
| Malawi | 0.063 | 114 | 5201 | 0.71 | 0.000643 |  |  |  |  |  |
| Mauritania | 0.139 | 90 | 5292 | 2.49 | 0.00202 |  |  |  |  |  |
| Yemen | 0.094 | 54 | 2092 | 3.10 | 2.22e-14 |  |  |  |  |  |
| Mean | 0.080 | 118 |  | 1.25 |  | 0.060 | 120 |  | 0.96 |  |
| Spain | 0.314 | 26 | 260692 | 5.66 | 0.000284 | 0.257 | 22 | 28822 | 4.91 | 2.34e-14 |
| UK | 0.137 | 51 | 307020 | 4.58 | 6.53e-08 | 0.208 | 30 | 41678 | 5.25 | 2.59e-12 |
| Italy | 0.185 | 32 | 241959 | 4.45 | 9.39e-05 | 0.188 | 29 | 35600 | 4.81 | 4.03e-13 |
| New York | 0.204 | 17 | 349598 | 3.91 | 0.00192 | 0.282 | 19 | 31682 | 5.82 | 0.000259 |
| Germany | 0.236 | 37 | 153489 | 3.40 | 0.00192 | 0.184 | 27 | 8818 | 3.44 | 0.000342 |
| France | 0.185 | 41 | 104231 | 2.17 | 0.00422 | 0.250 | 46 | 28161 | 4.05 | 0.000461 |
| Mean | 0.210 | 34 |  | 4.00 |  | 0.228 | 29 |  | 4.72 |  |

### Test of the Third Epidemic Law with Data of Multiple Epidemic Waves

We divide an epidemic system with multiple waves into three stages: The first stage is the exponential growth phase of the first wave that spreads under the natural environmental and normal societal conditions (Figs. 4 and 6). The second stage is affected by mitigation measures but a new normal is still developing (Fig. 6). When the new normal is developed, the epidemic enters the third stage that is characterized by a symmetric link between two consecutive waves or a symmetric wave with *m* = 1.

In what follows, we compare Eqs. (27a) and (28) with the COVID-19 data of the U.S. and Australia, which represent two different patterns under different mitigation measures.

### Data of Australia

Fig. 6 shows that Australia has experienced two complete epidemic waves with endemics. The first wave is asymmetric and broke out under the natural environmental and normal societal conditions. The second wave connects the first wave with a U-shaped symmetric link so the infection rate *µ*_1_/*m*_1_ in the first decay phase is equal to the infection rate *µ*_2_ in the second growth phase. This implies that the new normal condition has been developed since the first peak. Under the new normal condition, the second wave follows the classic logistic model and thus *m*_2_ = 1.

Based on these conditions, Eq. (28) is specified as:

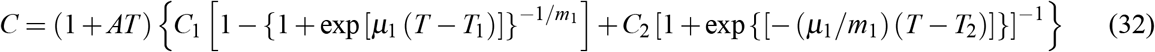

where *µ*_1_, *T*_1_, *C*_1_, *m*_1_, *T*_2_, *C*_2_ and *A* are fit parameters. Fitting Eq. (32) to the data in Fig. 9(a) results in:

**Figure 9:**
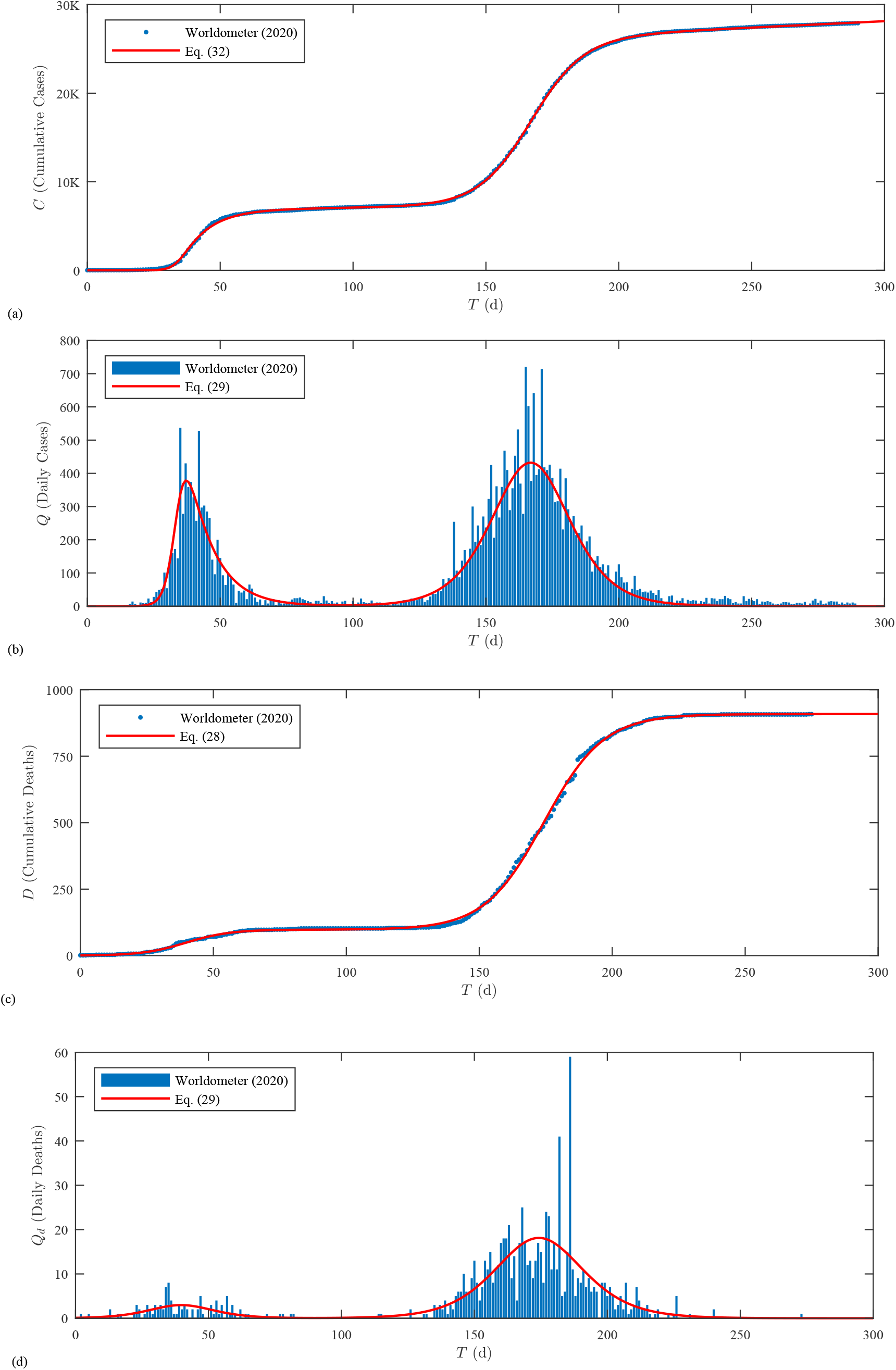
Test of the third epidemic law with data of COVID-19 in Australia: (a) cumulative confirmed cases; (b) daily confirmed cases; (c) cumulative deaths; (d) daily deaths.

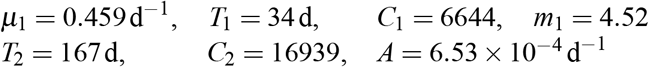

which yields the solid line in Fig. 9(a) with a determination coefficient *R*^2^ = 0.9999, meaning excellent agreement. Note that the number of cumulative cases in Australia increases linearly as *T* becomes very large until herd immunity is achieved, which is further explained with the daily cases.

The epidemic curve of daily cases from Eq. (29), together with the data, is plotted in Fig. 9(b). At *t* → ∞, Eq. (30) gives *Q*(∞) =(6.53 x 10 ^−4^) (6644 + 16939) = 15 cases/d, which implies that COVID-19 in Australia is controllable even without vaccines.

As for the death waves, we directly fit Eq. (28) to the cumulative death data in Fig. 9(c), resulting in

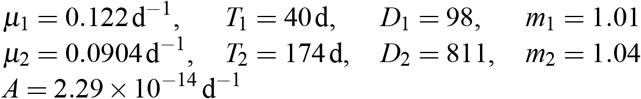

which yields the solid line in Fig. 9(c) with a determination coefficient *R*^2^ = 0.9996. The death epidemic curve from Eq. (29) and the data are then compared in Fig. 9(d). At *T*→∞, Eq. (30) gives *Q*_*d*_(∞)= 0, implying that COVID-19 in Australia has been controlled. The total deaths will be *D*(∞) = *D*_1_ + *D*_2_ = 909.

Generally, the values of *µ* for deaths are less than those for cases, implying the death grows slower than that of cases. Second, the natural inflection times *T*_*k*_ are comparable with those for cases, implying the death pattern follows that of cases. Third, both values of *m* for deaths are close to 1, implying that the COVID-19 death in Australia grows almost naturally and can be well described by a double-logistic model.

### Data of the U.S

The COVID-19 data in the U.S. are plotted in Fig. 10 that is different from Fig. 9 for Australia, because it does not have an endemic phase and two consecutive waves have a time overlap. Furthermore, a new normal condition (the third stage) with constant mitigation measures has not been developed because each state applied different measures in different times. For such a case, we fit Eq. (27a) to wave data one by one.

**Figure 10:**
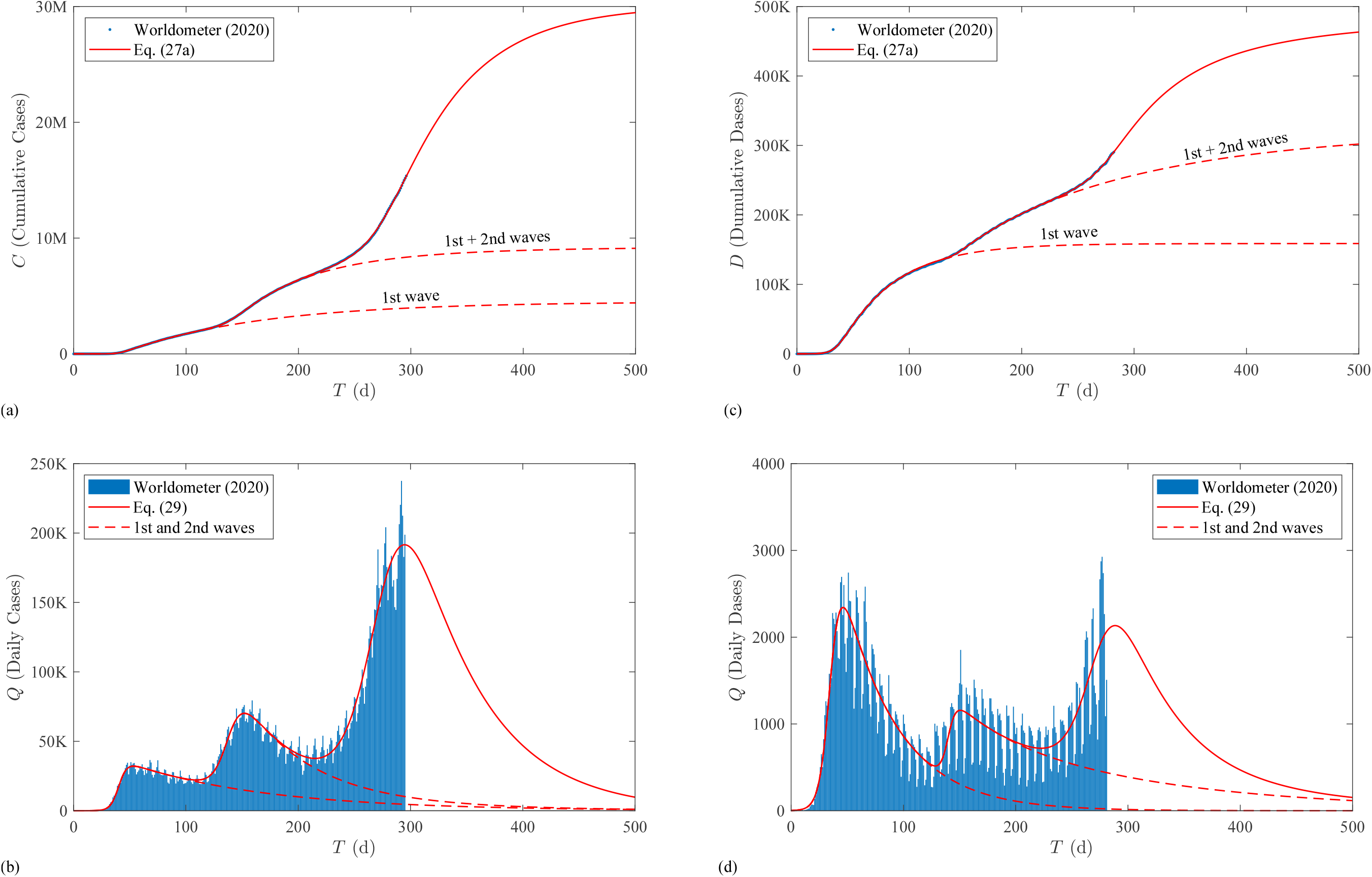
Test of the third epidemic law with data of COVID-19 in the U.S.: (a) cumulative confirmed cases; (b) daily confirmed cases; (c) cumulative deaths; (d) daily deaths.

We first apply Eq. (27a) to the data until *T* = 100 d in Fig. 10(b) for the first wave, namely,

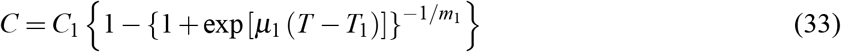

resulting in *µ*_1_ = 0.270 d^−1^, *T*_1_ = 40 d, *C*_1_ = 4.51 x10^6^, and *m*_1_ = 33 that means the first wave is significantly flattened. The resulting cumulative and daily cases equations are plotted in Figs. 10(a) and 10(b), respectively, with dashed lines.

We next add the second wave to the first wave and fit the data until *T* = 200 d in Fig. 10(b) for the second wave parameters, resulting in *µ*_2_ = 0.146 d^−1^, *T*_2_ = 138 d, *C*_2_ = 4.72×10^6^, and *m*_2_ = 8.60. The resulting cumulative and daily cases equations are also plotted in Fig. 10(a) and 10(b) with dashed lines. We see that *µ*_2_< *µ*_1_, *T*_2_−*T*_1_ = 98 d > *T*_1_ = 40 d and *m*_2_ > 1. Thus, the second wave is further suppressed.

We then add the third wave to the second one. Because the third wave has not yet completed in Fig. 10(b), we assume *m*_3_ = 4 based on the values in Table 1 for developed countries and the first wave of Australia. The other model parameters are then fitted as *µ*_3_ = 0.0647 d^−1^, *T*_3_ = 274 d, and *C*_3_ = 2.09 x 10^7^. We see that though *µ*_3_< *µ*_2_, *T*_3_ −*T*_2_ = 174 d > *T*_2_− *T*_1_ = 98 d, which suppress the third wave, the value of *C*_3_ is one order of magnitude larger than *C*_1_ and *C*_2_ and magnifies the daily cases significantly according to Eq. (10b). Therefore, the current mitigation measures in the U.S. cannot further suppress the COVID-19 transmission. The only way for the U.S. to return to normal is to vaccinate a large part of the population, as stated in Eq. (6), which then reduces the infection potential in Eq. (10b).

If without a fourth wave, the total cases in the U.S. will be *C*(∞)= *C*_1_ + *C*_2_ + *C*_3_ = 30M, which is approximately 10% of the U.S. population.

Fig. 10(d) shows that the death waves follow the case waves. Similarly, we can find the wave parameters as follows:

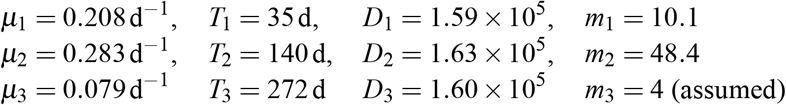

which plot Eqs. (27a) and (29) with *A* = 0 in Figs. 10(c) and 10(d), respectively. The total deaths in the U.S. will be *D* (∞)= *D*_1_ + *D*_2_ + *D*_3_ = 482K. The terminal case fatality rate in the U.S. is then approximately 482000/(30×10^6^) = 1. 61%.

Briefly, the data of the U.S. and Australia validate the third epidemic law and its corollary. Yet, further tests with the whole world data, including the U.S. states data, are needed in future studies. Besides, To incorporate the proposed theory into the classic SIR model is also a research need, which is briefed below.

### Future Research: SIR Model with Intervention Measures

Eq. (11a) describes the production of daily new cases, which can be used to improve the classic SIR model proposed by Kermack and McKendrick (1927), who further divided *C(T)* in Eq. (5) into active infectious cases *I(T)* and removed cases *R(T)* The SIR model reads:

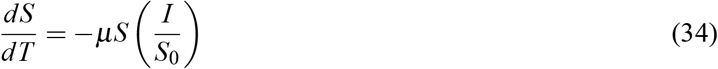

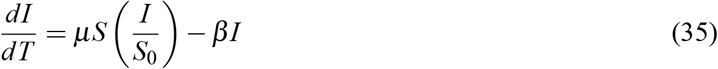

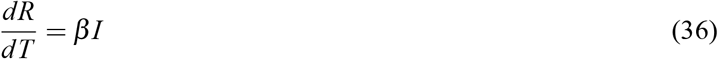

where *S*_0_ = initial susceptible or the system population if *P*_*h*_ = *P*_*v*_ = 0 in Eqs. (5) and (6); *I/S*_0_ = probability that susceptible meet infected individuals; and *β* = recovery rate. Eq. (34) is the daily decrease of susceptible; Eq. (35) is the daily change of active cases; and Eq. (36) is the daily removed cases including deaths and recovered.

If intervention measures are applied, according to Eq. (11a), the infection rate *µ* becomes the effective infection rate *µ/m*, and the probability that susceptible meet infected individuals becomes [1 −(1 −*I/S*_0_)^*m*^]. Therefore, Eqs. (34) and (35) are modified as:

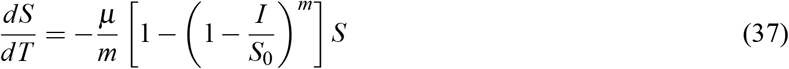

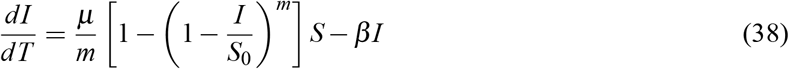

These two equations reduce to Eqs. (34) and (35) at the initial growth where *I* ≪*S*_0_ and at *m* = 1 if without intervention measures. The detailed study of intervention measures on the SIR model through Eqs. (36)-(38) is a future research need in epidemiology.

## Conclusions

Based on the world COVID-19 data, we divided an epidemic curve into three phases: a growth phase, a decay phase, and an endemic phase. We then discovered two simple epidemic laws inductively and deductively to describe the three phases mathematically. Furthermore, we extended the two laws to an epidemic with multiple epidemic waves to obtain a third epidemic law. Specifically, we reach the following conclusions:

- *The first epidemic law* states that given an isolated epidemic system with human interventions, if the epidemic decays out asymptotically without an endemic phase, the curve of cumulative cases is a sigmoid curve (Fig. 3) described by the complement of the generalized Fermi function [Eq. (17a)]. As a corollary, this law leads to an epidemic curve that grows and decays both exponentially, but the decay phase is flattened by intervention mitigation measures [Eq. (19b) and Fig. 4]. The classic logistic law is a special case under constant environmental and normal societal conditions.
- *The second epidemic law* combines the first law and the endemic phase [Eq. (24b)]. It states that if an epidemic has an endemic phase, the number of cumulative cases increases linearly as time tends to infinity [Fig. 5(a)]. As a corollary, this law states that the number of daily new cases grows exponentially, decays exponentially, but tends to a constant in the endemic phase [Fig. 5(b)]. If the endemic constant is small, the epidemic is controllable with mitigation measures even without vaccines; otherwise, it is out of control if without vaccines.
- *The third epidemic law* is a superposition principle for an epidemic system with multiple epidemic waves. It states that: (1) If there is not an endemic phase between two consecutive waves, then the resultant number of cumulative cases is a superposition of the first epidemic law [Eq. (27a)]. (2) If an epidemic has an endemic phase even after multiple epidemic waves, then the resultant number of cumulative cases is a superposition of the second epidemic law [Eq. (28)]. Similarly, an corollary can be deduced for epidemic curves with multiple epidemic waves.
- The three epidemic laws are confirmed with the COVID-19 data of cumulative cases from 18 countries including undeveloped, developing and developed countries (Figs. 7 to 10). The three corollaries agree with the time series data of daily new cases. Besides, the three laws and the three corollaries are validated with the COVID-19 death data because of similarities of epidemic curves between daily new cases and daily deaths (Figs. 7 to 10). To further validate the three epidemic laws, extensive tests with other COVID-19 data, including the U.S. states data, and other epidemic data are required in future studies.
- Finally, we paved the way for future research to incorporate the proposed theory into the classic SIR model.

## Data Availability

All data referred to in the manuscript are from Worldometer.

https://www.worldometers.info/coronavirus/

## References

Fermi, E., Pasta, J. R., and Ulam, S. (1955). Studies of nonlinear problems. I. Report LA-1940. Los Alamos: Los Alamos Scientific Laboratory.

Gompertz, B. (1825). “On the nature of the function expressive of the law of human mortality, and on a new mode of determining the value of life contingencies.” Philos. Trans. R. Soc. Lond. 115, 513–583.

Guo, J. (1988). “Vertical distributions of velocity and sediment concentration in wide open channels.” Journal of Taiyuan University of Technology, 19(2), 42-49 (in Chinese). http://en.cnki.com.cn/Article_en/CJFDTotal-TYGY198802005.htm

Kermack, W. O., and McKendrick, A. G. (1927). “A Contribution to the Mathematical Theory of Epidemics.… Proc. Roy. Soc. Lond. A 115, 700–721.

Korteweg D. J., and de Vries G. (1895). “On the change of form of long waves advancing in a rectangular canal, and on a new type of long stationary waves.” Philos Mag (Ser 5) 39:422–443.

Richards, F. J. (1959). “A flexible growth function for empirical Use.” Journal of Experimental Botany. 10 (2): 290–300.

Wazwaz, A. M. (2009). Solitary Waves Theory. In: Partial Differential Equations and Solitary Waves Theory. Nonlinear Physical Science. Springer, Berlin, Heidelberg. https://doi.org/10.1007/978-3-642-00251-9_12/.

WHO (2020). https://www.who.int/.

Worldometer (2020). COVID-19 Coronavirus Pandemic: https://www.worldometers.info/coronavirus/.

